## Supplement for "Clinical outcomes and cost-effectiveness of COVID-19 vaccination in South Africa"

**Supplementary Tables** 14

**Supplementary Table 1. Model-projected 360-day clinical outcomes and costs of a reference COVID-19 vaccination program compared with no vaccination program in South Africa.** 14

**Supplementary Table 2. Clinical and economic outcomes of different COVID-19 vaccination program strategies of vaccine supply and vaccination pace under scenarios of slow and rapid epidemic growth in South Africa.** 16

**Supplementary Table 3. Additional one-way sensitivity analyses, clinical and economic outcomes of a reference COVID-19 vaccination program compared with no vaccination program in South Africa.** 17

**Supplementary Table 4. Clinical and economic outcomes of different COVID-19 vaccination program strategies of vaccine supply and vaccination pace under different scenarios of epidemic growth in South Africa: applying lower costs of hospital and ICU care from Edoka et al.**[3] 19

**Supplementary Table 5. Additional model input parameters.** 22

**Supplementary Methods** 24

**Overview** 24

**Health states** 25

**Transmission** 27

**Disease natural history** 28

**Life expectancy and years-of-life lost** 29

**Vaccine effectiveness and outcomes** 30

**Costs of vaccination, hospital ward care, and ICU care** 32

**Model validation** 33

**Supplementary References** 34

**Supplementary Figures**

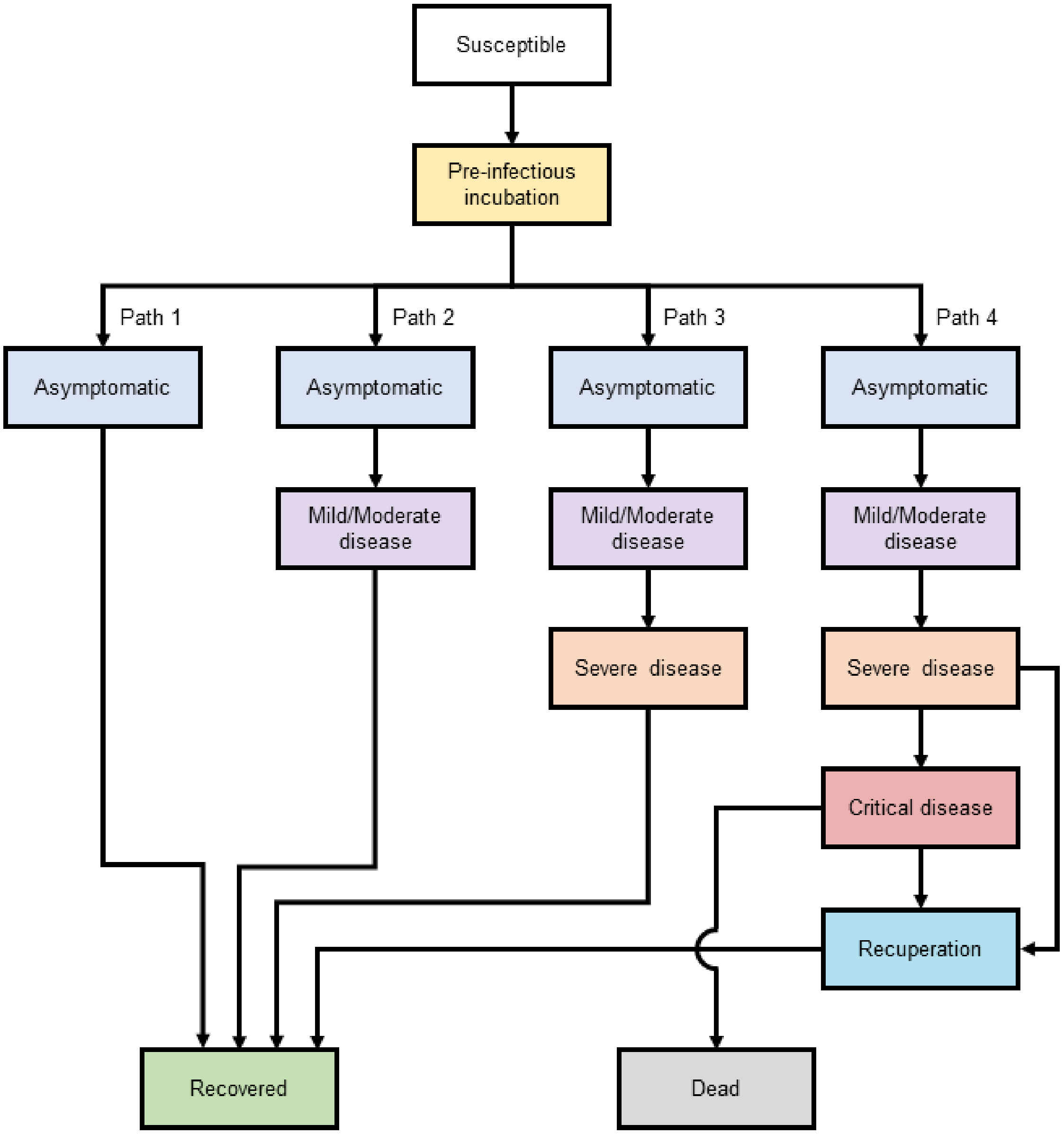

**Supplementary Figure 1. Health states and disease paths in the Clinical and Economic Analysis of COVID-19 Interventions (CEACOV) model.** Mild/moderate COVID-19 disease is that which does not prompt hospitalization. Severe disease prompts hospitalization if hospital (non-ICU) beds are available. Critical disease prompts ICU admission if ICU beds are available.

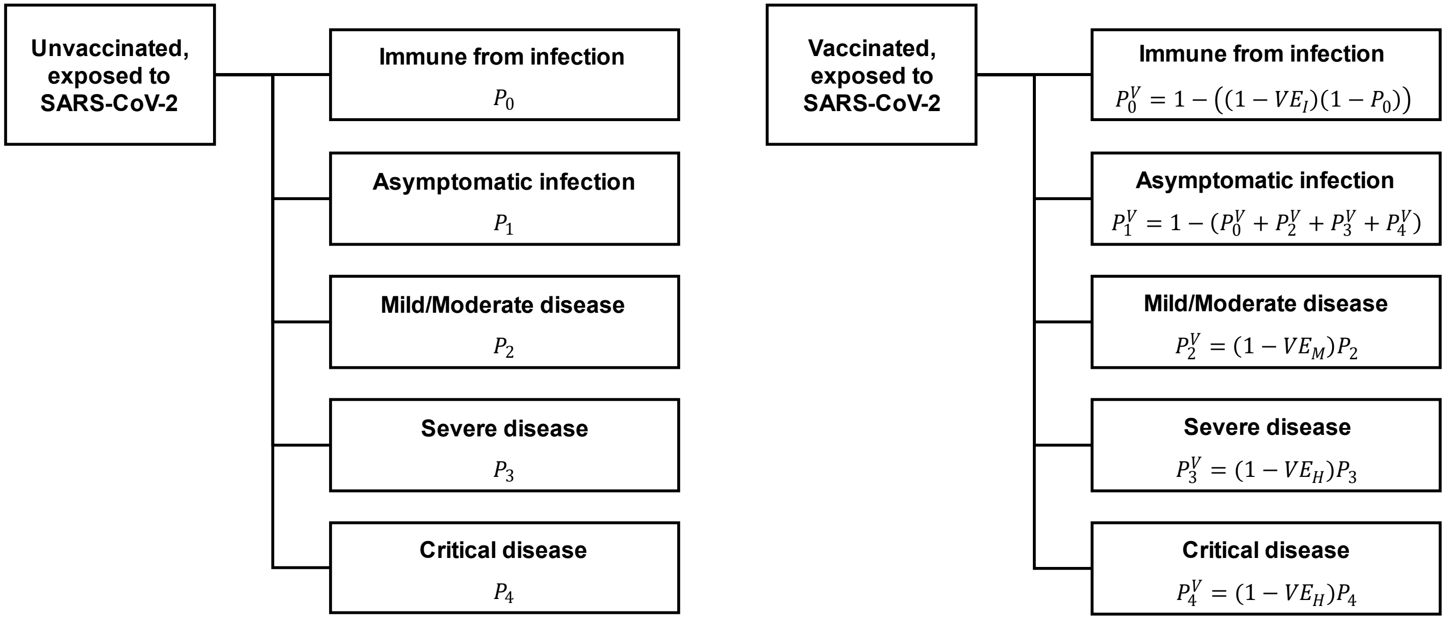

**Age-dependent disease path probabilities**

$P_0$: Probability immune from infection
$P_1$: Probability of developing asymptomatic infection if exposed
$P_2$: Probability of developing mild/moderate disease if exposed
$P_3$: Probability of developing severe disease if exposed
$P_4$: Probability of developing critical disease if exposed

**Vaccine effectiveness measures**

$VE_I$: Vaccine effectiveness in preventing infection
$VE_M$: Vaccine effectiveness in preventing mild/moderate disease
$VE_H$: Vaccine effectiveness in preventing severe or critical disease requiring hospitalization

**Supplementary Figure 2. Relationship between vaccine effectiveness and disease path probabilities among those vaccinated.** The tree diagram on the left depicts the probability of different disease paths among unvaccinated individuals exposed to SARS-CoV-2, while the diagram on the right depicts the probability of these same disease paths among vaccinated individuals. Our analysis considers three measures of vaccine effectiveness: effectiveness in preventing SARS-CoV-2 infection ($VE_I$), in preventing mild/moderate COVID-19 disease ($VE_M$), and in preventing severe or critical COVID-19 disease that would prompt hospitalization ($VE_H$).

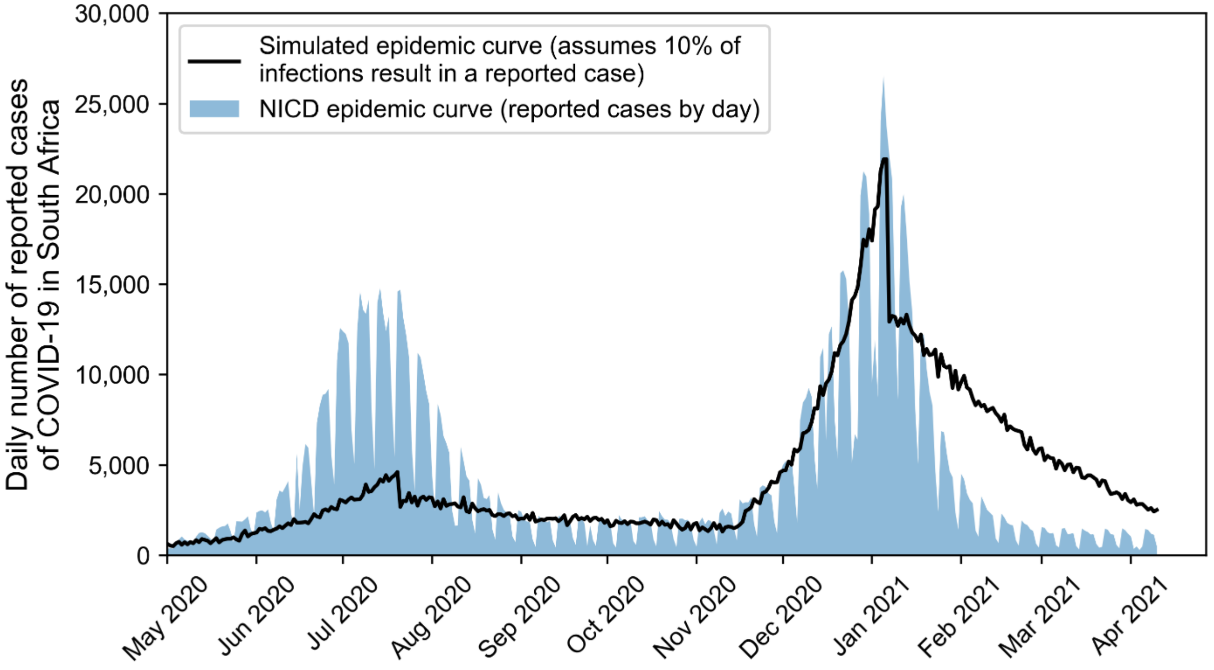

**Supplementary Figure 3. Model validation: comparison of "reported" model-projected SARS-CoV-2 infections and reported cases to South Africa's National Institute for Communicable Diseases.** The black line represents 0.1 times the daily number of model-projected SARS-CoV-2 infections (both reported and unreported) in South Africa, reflecting an assumption that 10% of infections are reported. Accordingly, the actual model-projected total number of infections was 10 times that shown by the black line. This modeled scenario starts with 0.1% prevalence of active SARS-CoV-2 infection and 0% prevalence of prior immunity (analogous to South Africa in early 2020). The blue bars represent COVID-19 cases reported to South Africa's National Institute for Communicable Diseases (NICD).[1,2]

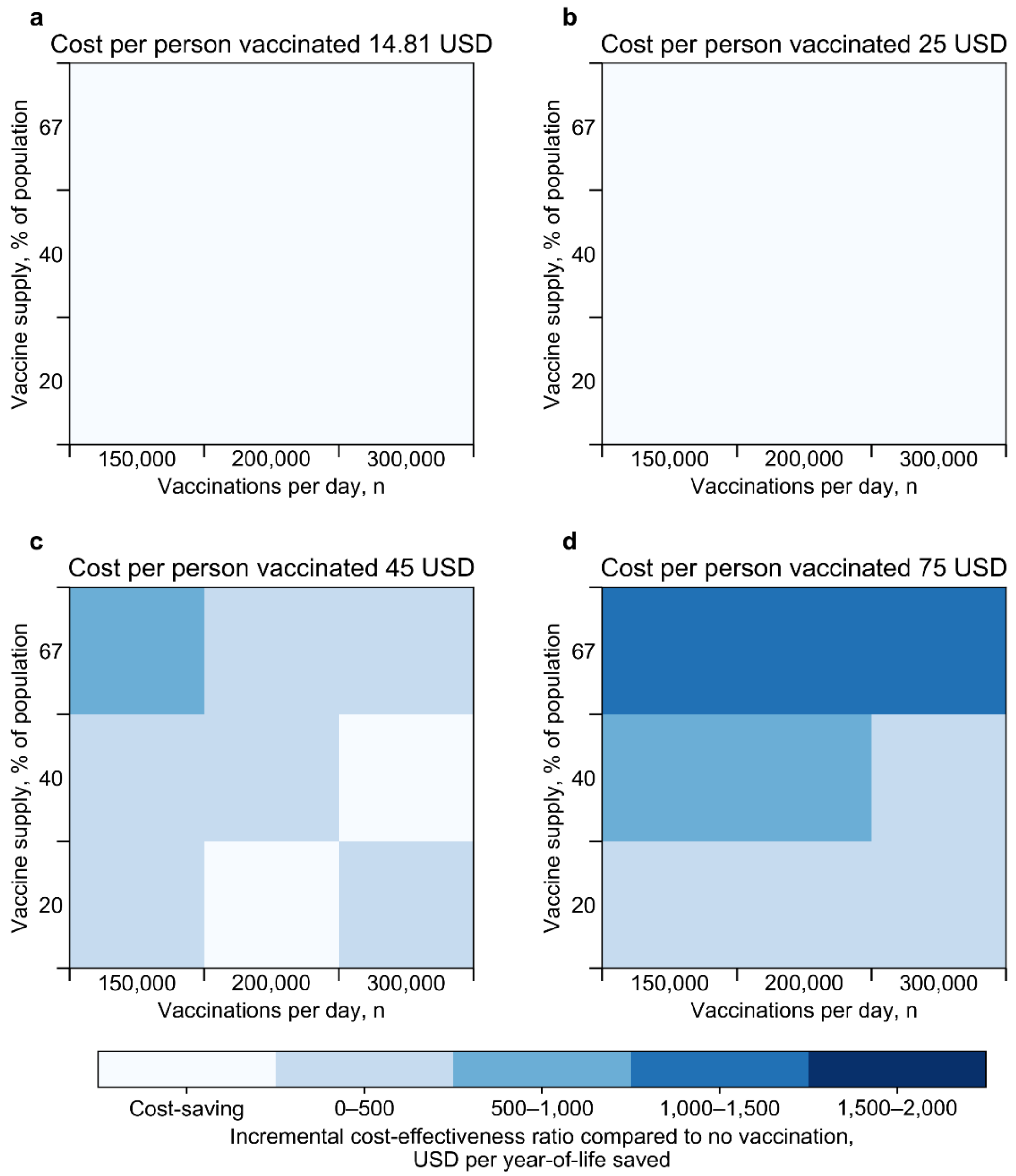

**Supplementary Figure 4. Multi-way sensitivity analysis of vaccine supply, vaccination pace, and vaccination cost: incremental cost-effectiveness ratio of vaccination program compared with no vaccination.** Within each panel, the smaller boxes are colored according to the incremental cost-effectiveness ratio (ICER). The lightest color represents scenarios in which a reference vaccination program (vaccine supply sufficient for 67% of South Africa's population, pace 150,000 vaccinations per day) is cost-saving compared with no vaccination program, meaning that it results in clinical benefit and reduces overall health care costs. The darker colors reflect increasing ICERs, whereby a reference vaccination program, compared with no vaccination program, results in both clinical benefit and higher overall health care costs. The ICER is the model-generated difference in costs divided by the difference in years-of-life between a reference vaccination program and no vaccination program.

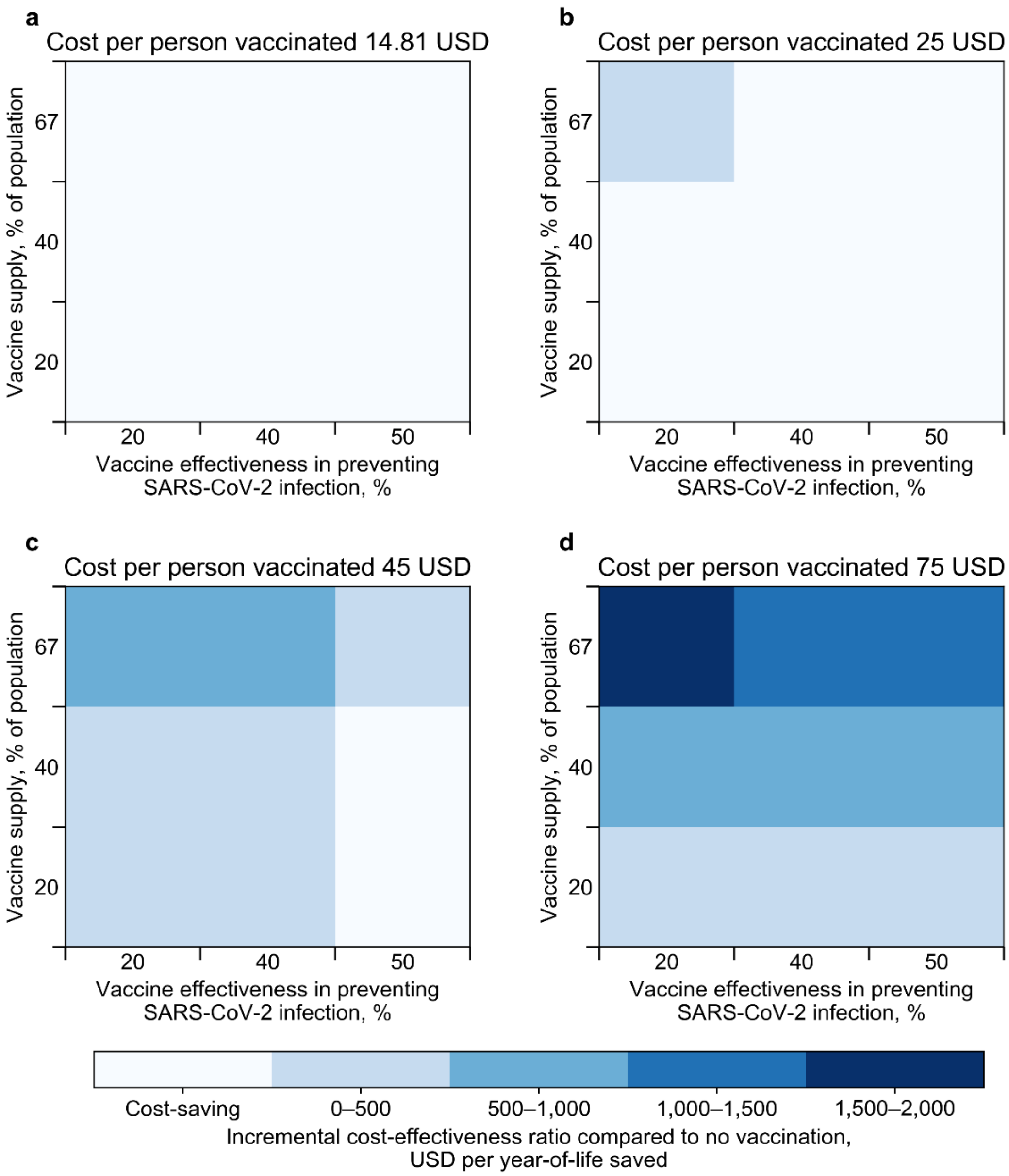

**Supplementary Figure 5. Multi-way sensitivity analysis of vaccine supply, vaccine effectiveness against infection, and vaccination cost: incremental cost-effectiveness ratio of vaccination program compared with no vaccination.** Within each panel, the smaller boxes are colored according to the incremental cost-effectiveness ratio (ICER). The lightest color represents scenarios in which a reference vaccination program (vaccine supply sufficient for 67% of South Africa's population, pace 150,000 vaccinations per day) is cost-saving compared with no vaccination program, meaning that it results in clinical benefit and reduces overall health care costs. The darker colors reflect increasing ICERs, whereby a reference vaccination program, compared with no vaccination program, results in both clinical benefit and higher overall health care costs. The ICER is the model-generated difference in costs divided by the difference in years-of-life between a reference vaccination program and no vaccination program.

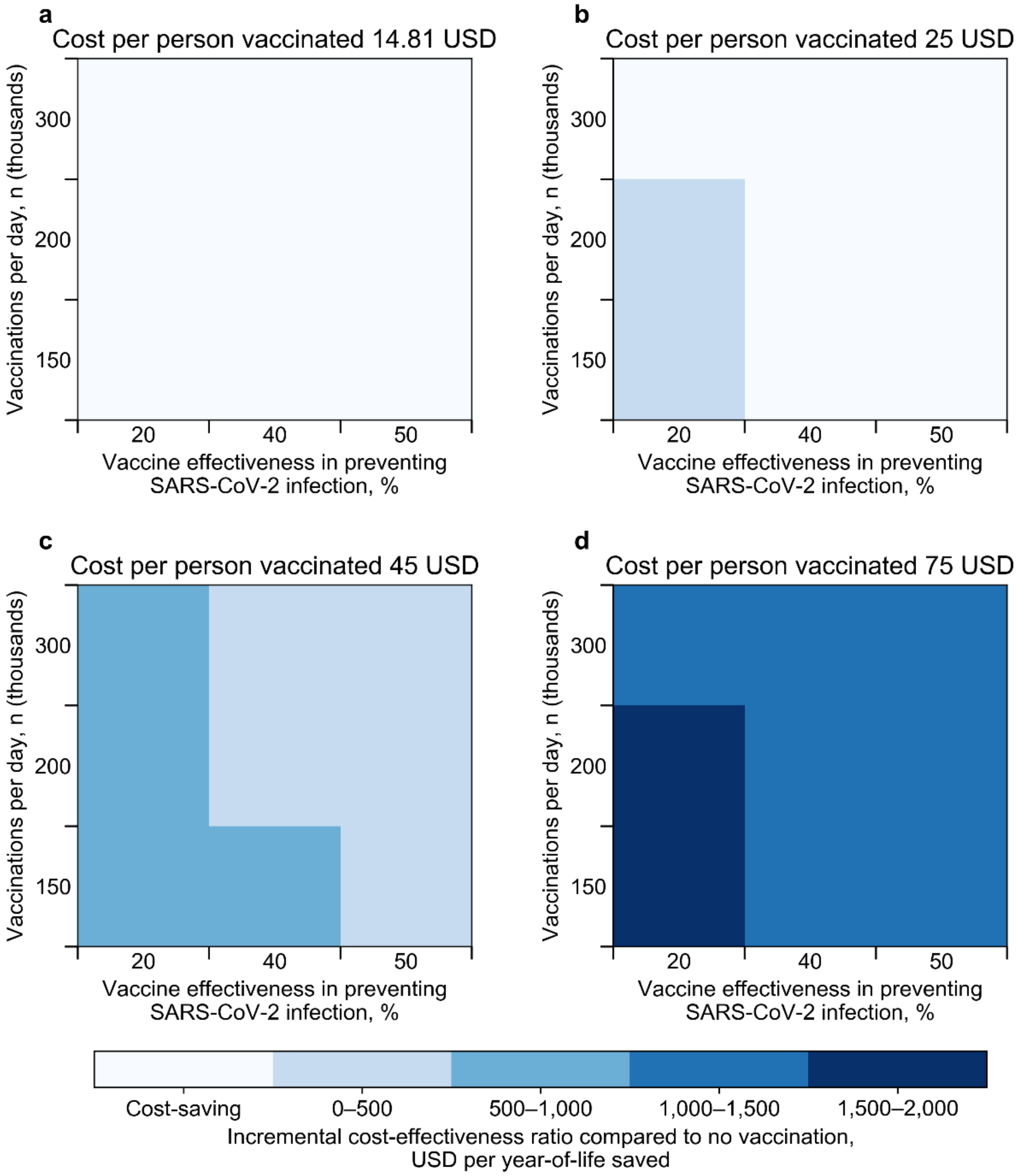

**Supplementary Figure 6. Multi-way sensitivity analysis of vaccination pace, vaccine effectiveness against infection, and vaccination cost: incremental cost-effectiveness ratio of vaccination program compared with no vaccination.** Within each panel, the smaller boxes are colored according to the incremental cost-effectiveness ratio (ICER). The lightest color represents scenarios in which a reference vaccination program (vaccine supply sufficient for 67% of South Africa's population, pace 150,000 vaccinations per day) is cost-saving compared with no vaccination program, meaning that it results in clinical benefit and reduces overall health care costs. The darker colors reflect increasing ICERs, whereby a reference vaccination program, compared with no vaccination program, results in both clinical benefit and higher overall health care costs. The ICER is the model-generated difference in costs divided by the difference in years-of-life between a reference vaccination program and no vaccination program.

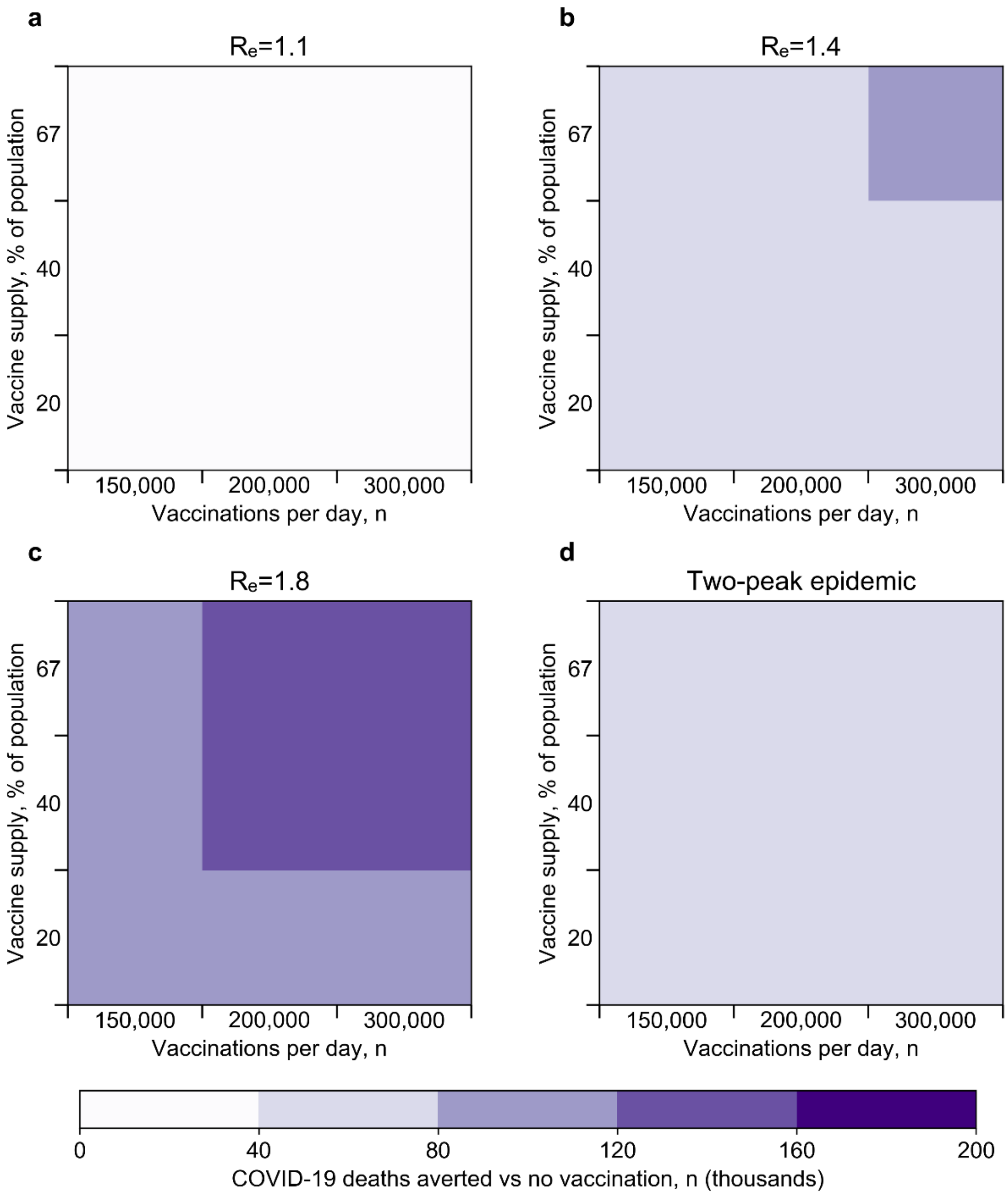

**Supplementary Figure 7. Multi-way sensitivity analysis of vaccine supply, vaccination pace, and effective reproduction number ($R_e$): number of COVID-19 deaths (in thousands) averted through a vaccination program compared with no vaccination.** Within each panel, the smaller boxes are colored according to the number of COVID-19 deaths averted over 360 days by a vaccination program compared with no vaccination program. In the analysis of an epidemic with two peaks, the basic reproduction number ($R_o$) alternates between low and high values over time, and the $R_e$ changes day-to-day as the epidemic and vaccination program progress and there are fewer susceptible individuals. For most of the simulation horizon, $R_o$ is 1.6 (equivalent to an initial $R_e$ of 1.1). However, during days 90-150 and 240-300 of the simulation, $R_o$ is increased to 2.6. This results in two epidemic waves with peak $R_e$ of approximately 1.4-1.5.

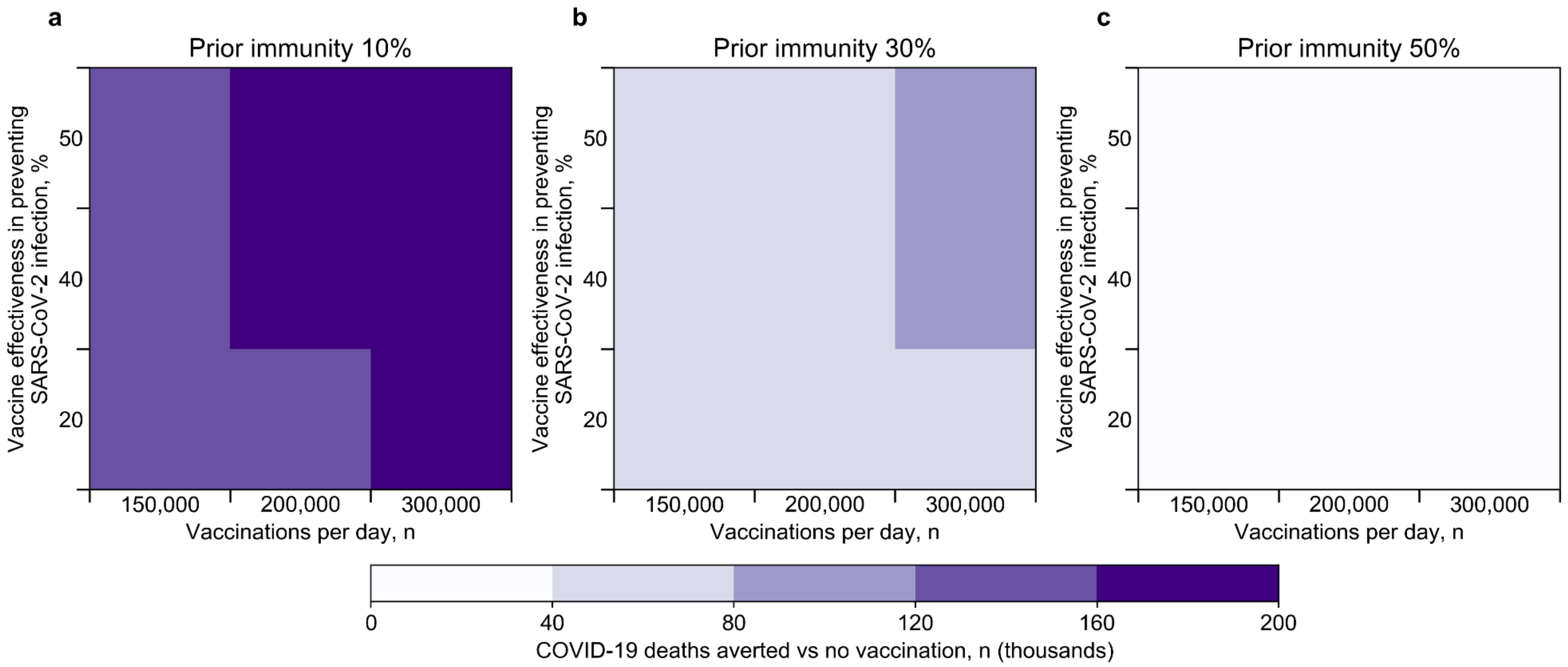

**Supplementary Figure 8. Multi-way sensitivity analysis of vaccine effectiveness against infection, vaccination pace, and prevalence of prior immunity to SARS-CoV-2: number of COVID-19 deaths (in thousands) averted through a vaccination program compared with no vaccination.** Within each panel, the smaller boxes are colored according to the number of COVID-19 deaths averted over 360 days by a vaccination program compared with no vaccination program.

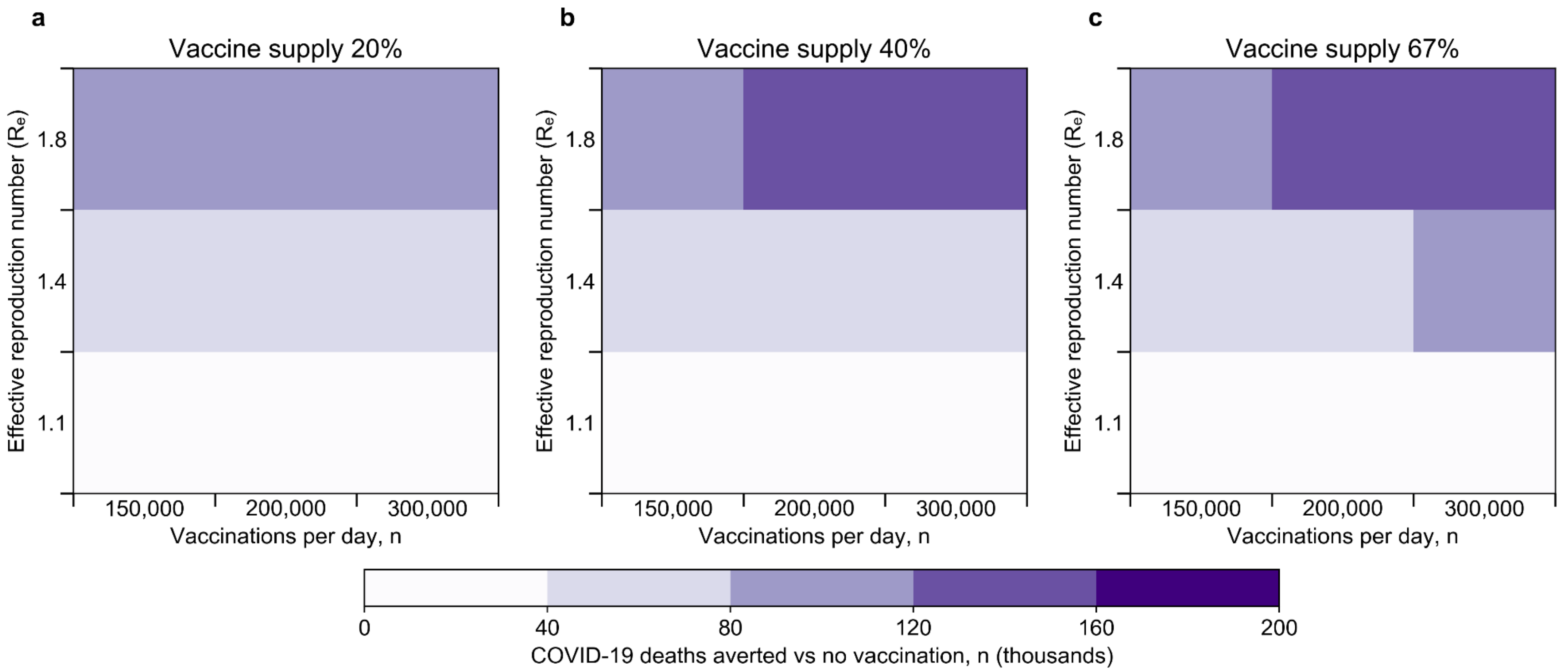

**Supplementary Figure 9. Multi-way sensitivity analysis of effective reproduction number ($R_e$), vaccination pace, and vaccine supply: number of COVID-19 deaths averted by vaccination program.** Within the heatmap, the smaller boxes are colored according to the number of COVID-19 deaths averted over 360 days by a vaccination program compared with no vaccination program.

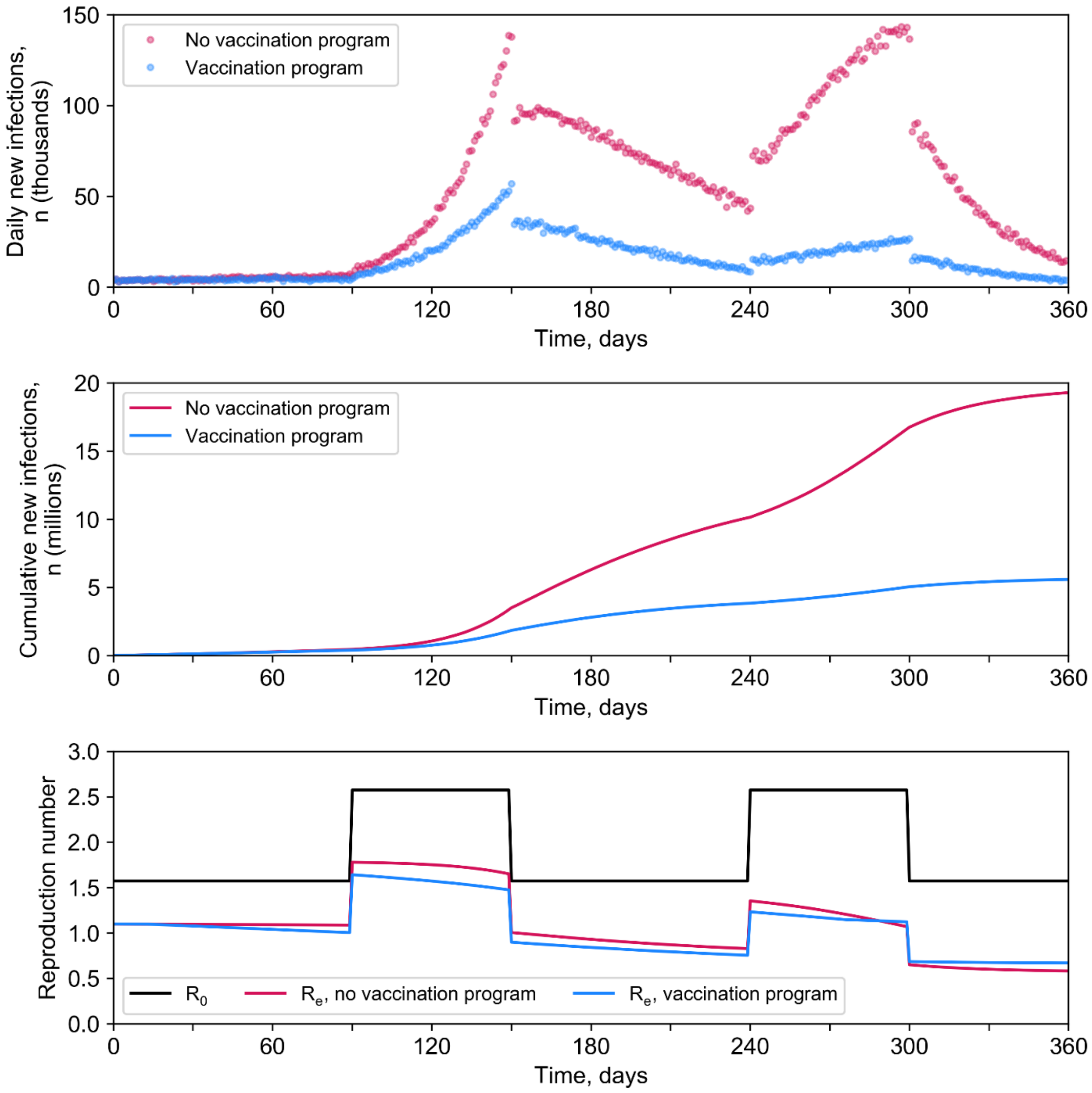

**Supplementary Figure 10. Model projection of epidemic with varying effective reproduction number ($R_e$).** We evaluated a scenario in which there were episodic surges above a lower background basic reproduction number ($R_o$). We did this by alternating the $R_o$ between low and high values over time. This results in the $R_e$ changing day-to-day as the epidemic and vaccination program progress and there are fewer susceptible individuals. For most of the simulation horizon, $R_o$ is 1.6 (equivalent to an initial $R_e$ of 1.1). However, during days 90-150 and 240-300 of the simulation, $R_o$ is increased to 2.6. This results in two epidemic waves with peak $R_e$ of approximately 1.4-1.5.

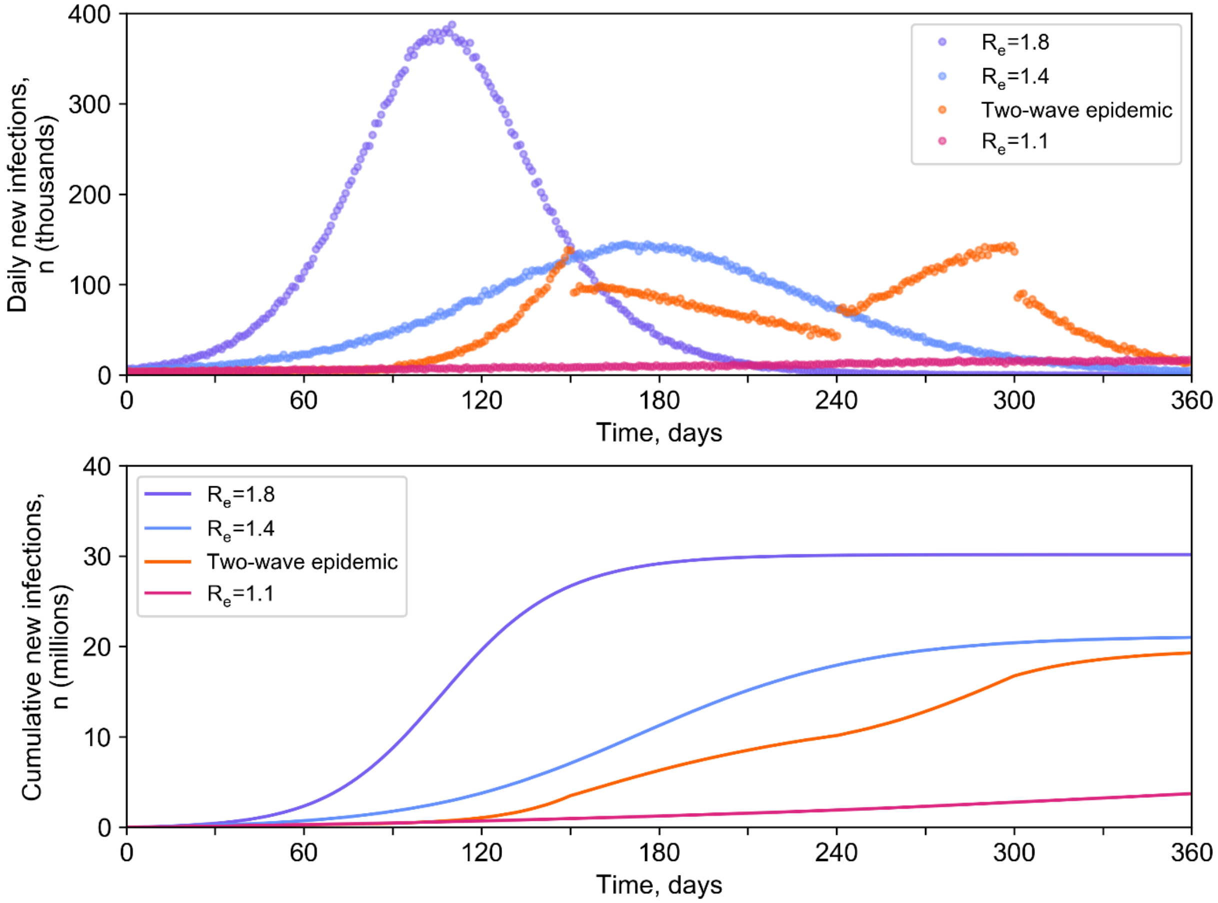

**Supplementary Figure 11. Modeled scenarios of epidemic growth in the absence of vaccines.** We evaluated four epidemic growth scenarios: three in which $R_o$ remains constant over time, resulting in an initial $R_e$ of 1.1, 1.4, or 1.8; and one in which $R_o$ is modulated over time to produce an epidemic with two waves

### Supplementary Tables

**Supplementary Table 1. Model-projected 360-day clinical outcomes and costs of a reference COVID-19 vaccination program compared with no vaccination program in South Africa.**

| Parameter | No vaccination program | Vaccination program[a], (% reduction compared with no vaccination program) |
|---|---|---|
| Cumulative SARS-CoV-2 infections, n | | |
| Ages <20 years | 7,695,800 | 4,355,700 (43%) |
| Ages 20-59 years | 11,410,000 | 5,402,600 (53%) |
| Ages ≥60 years | 1,906,300 | 826,700 (57%) |
| Overall | 21,012,100 | 10,585,100 (50%) |
| Cumulative COVID-19 deaths, n | | |
| Ages <20 years | 500 | 200 (61%) |
| Ages 20-59 years | 37,500 | 6,000 (84%) |
| Ages ≥60 years | 51,300 | 8,500 (83%) |
| Overall | 89,300 | 14,700 (84%) |
| Resource use | | |
| Hospital bed-days, n | 2,981,400 | 984,900 (67%) |
| ICU bed-days, n | 746,300 | 344,700 (54%) |
| Peak daily hospital occupancy, n | 21,100 | 6,400 (70%) |
| Peak daily ICU occupancy, n | 3,300[b] | 2,200 (33%) |
| Health care costs, USD | | |
| Hospital care | 460,063,200 | 151,975,800 (67%) |
| ICU care | 1,306,793,100 | 603,620,800 (54%) |
| Vaccinations | -- | 583,206,900 |
| Cost-effectiveness | | |
| Years-of-life lost, n | 1,558,700 | 259,600 (83%) |
| Total cost, USD | 1,766,856,200 | 1,338,803,500 (24%) |
| ICER compared with no vaccination program, USD per year-of-life saved | -- | Cost-saving[c] |

ICU: intensive care unit. USD: United States dollars. ICER: incremental cost-effectiveness ratio.

[a]In the reference vaccination program, there is sufficient vaccine supply for 67% of South Africa's population, and 150,000 vaccinations are administered per day. Those aged ≥60 years are prioritized. Uptake (acceptance) of vaccine is 67% across all ages.

[b]Maximum ICU capacity is reached.

[c]Because the vaccination strategy results in fewer years-of-life lost (greater clinical benefit) and lower health care costs, it is cost-saving and no ICER is displayed.

**Supplementary Table 2. Clinical and economic outcomes of different COVID-19 vaccination program strategies of vaccine supply and vaccination pace under scenarios of slow and rapid epidemic growth in South Africa.**

| Scenario and Vaccination Strategy | Cumulative SARS-CoV-2 infections | Cumulative COVID-19 deaths | Years-of-life lost | Health care costs, USD | ICER, USD per year-of-life saved[a] |
|---|---|---|---|---|---|
| $R_e$ = 1.1[b] | | | | | |
| No vaccination | 3,719,500 | 8,200 | 123,500 | 382,176,900 | -- |
| Vaccine supply 40%, pace 300,000 vaccinations per day | 860,900 | 1,300 | 24,500 | 417,756,900 | 360 |
| Vaccine supply 40%, pace 150,000 vaccinations per day | 1,144,800 | 1,400 | 24,300 | 449,386,900 | Dominated |
| Vaccine supply 67%, pace 300,000 vaccinations per day | 718,900 | 1,300 | 21,700 | 652,819,400 | 85,290 |
| Vaccine supply 67%, pace 150,000 vaccinations per day | 1,079,100 | 1,500 | 25,400 | 681,669,900 | Dominated |
| $R_e$ = 1.8 | | | | | |
| Vaccine supply 40%, pace 300,000 vaccinations per day | 21,260,400 | 38,800 | 695,700 | 1,541,112,600 | -- |
| Vaccine supply 40%, pace 150,000 vaccinations per day | 24,268,200 | 69,500 | 1,410,500 | 1,596,237,800 | Dominated |
| Vaccine supply 67%, pace 300,000 vaccinations per day | 18,511,000 | 33,800 | 593,600 | 1,659,492,700 | 1,160 |
| No vaccination | 30,173,000 | 179,300 | 3,360,700 | 1,673,830,400 | Dominated |
| Vaccine supply 67%, pace 150,000 vaccinations per day | 24,217,300 | 68,900 | 1,403,000 | 1,803,189,900 | Dominated |

USD: United States dollars. ICER: incremental cost-effectiveness ratio. $R_e$: effective reproduction number. Dominated: the strategy results in a higher ICER than that of a more clinically effective strategy, or the strategy results in less clinical benefit (more years-of-life lost) and higher health care costs than an alternative strategy.

[a]Within each $R_e$ scenario, vaccination strategies are ordered from lowest to highest cost per convention of cost-effectiveness analysis. ICERs are calculated compared to the next least expensive, non-dominated strategy. Displayed life-years and costs are rounded to the nearest hundred, while ICERs are calculated based on non-rounded life-years and costs and then rounded to the nearest ten.

[b]Because there are relatively few deaths in the $R_e$=1.1 scenario, the impact of changing vaccination program strategies is attenuated. Due to stochastic variation in simulation model results, and extrapolation from a simulated population of 1 million to South Africa's population of approximately 59 million, results indicate a slight increase in deaths when increasing vaccine supply while pace is maintained at 150,000 vaccinations per day. Also, due to differences in the ages of those who die, there can be discordance between number of deaths and number of years-of-life lost.

**Supplementary Table 3. Additional one-way sensitivity analyses, clinical and economic outcomes of a reference COVID-19 vaccination program compared with no vaccination program in South Africa.**

| Parameter / Value | SARS-CoV-2 infections averted, compared with no vaccination | COVID-19 deaths averted, compared with no vaccination | Years-of-life saved, compared with no vaccination | Change in health care costs, compared with no vaccination, USD | ICER, compared with no vaccination, USD per YLS[a] |
|---|---|---|---|---|---|
| Relative reduction in onward transmission rate among vaccinated individuals who become infected, % | | | | | |
| 0 (base case) | 10,427,000 | 74,600 | 1,299,100 | -428,052,700 | Cost-saving |
| 50 | 15,871,900 | 82,200 | 1,440,100 | -741,198,800 | Cost-saving |
| Cost of hospital and ICU care, relative to base case values | | | | | |
| 0.5x | 10,427,000 | 74,600 | 1,299,100 | 77,577,100 | 60 |
| 1.0x (base case) | 10,427,000 | 74,600 | 1,299,100 | -428,052,700 | Cost-saving |
| 2.0x | 10,427,000 | 74,600 | 1,299,100 | -1,439,312,400 | Cost-saving |
| Heath care costs considered | | | | | |
| Vaccination | 10,427,000 | 74,600 | 1,299,100 | 583,206,900 | 450 |
| Vaccination and hospital (non-ICU) care | 10,427,000 | 74,600 | 1,299,100 | 275,119,500 | 210 |
| Vaccination, hospital care, and ICU care (base case) | 10,427,000 | 74,600 | 1,299,100 | -428,052,700 | Cost-saving |
| Time to start of rollout, days | | | | | |
| 0 (base case) | 10,427,000 | 74,600 | 1,299,100 | -428,052,700 | Cost-saving |
| 30 | 8,395,100 | 72,900 | 1,287,000 | -243,551,400 | Cost-saving |
| 60 | 6,330,900 | 65,900 | 1,151,200 | 6,364,700 | 10 |
| Time to start of epidemic wave, days[b] | | | | | |
| 0 (base case) | 10,427,000 | 74,600 | 1,299,100 | -428,052,700 | Cost-saving |
| 30 | 12,382,700 | 78,100 | 1,401,300 | -592,758,700 | Cost-saving |
| 90[c] | 14,907,000 | 71,700 | 1,254,400 | -683,697,100 | Cost-saving |

USD: United States dollars. ICER: incremental cost-effectiveness ratio. YLS: year-of-life saved. $R_e$: effective reproduction number.

[a]In the reference vaccination program, there is sufficient vaccine supply for 67% of South Africa's population, and 150,000 vaccinations are administered per day. Those aged ≥60 years are prioritized. Uptake (acceptance) of vaccine is 67% across all ages. Displayed life-years and costs are rounded to the nearest hundred, while ICERs are calculated based on non-rounded life-years and costs and then rounded to the nearest ten. Cost-saving reflects more years-of-life saved (greater clinical benefit) and lower costs, and therefore ICERs are not displayed.

**[b]**In the analysis of a delayed epidemic wave, the basic reproduction number ($R_o$) is initially low but later assumes a higher value. Prior to the start of the epidemic wave, $R_o$ is 1.6 (equivalent to an initial $R_e$ of 1.1). After a set number of days, $R_o$ increases to 2.0, resulting in an epidemic wave with initial $R_e$ of approximately 1.4.

[c]When the epidemic wave is delayed by 90 days, the epidemic peak occurs more than 180 days into the simulation. Because our modeled time horizon only considers outcomes occurring through day 360, this leads to a small decrease in the number of infections and deaths that are recorded in the scenario without vaccines. As a result, the absolute number of deaths prevented by vaccines decreases slightly as the time to the start of the epidemic wave is increased from 30 to 90 days, even though a greater proportion of deaths are prevented.

**Supplementary Table 4. Clinical and economic outcomes of different COVID-19 vaccination program strategies of vaccine supply and vaccination pace under different scenarios of epidemic growth in South Africa: applying lower costs of hospital and ICU care from Edoka et al.[3]**

| Scenario and Vaccination Strategy | Cumulative SARS-CoV-2 infections | Cumulative COVID-19 deaths | Years-of-life lost | Health care costs, USD | ICER, USD per year-of-life saved[a] |
|---|---|---|---|---|---|
| Vaccine supply | | | | | |
| $R_e$ = 1.4 | | | | | |
| Vaccine supply 40% | 11,784,700 | 16,000 | 275,800 | 797,871,600 | -- |
| Vaccine supply 20% | 15,489,500 | 21,800 | 397,300 | 890,934,300 | Dominated |
| Vaccine supply 67% | 10,585,100 | 14,700 | 259,600 | 993,638,400 | Dominated |
| No vaccination | 21,012,100 | 89,300 | 1,558,700 | 1,005,281,800 | Dominated |
| Vaccine supply 80%[b] | 10,410,000 | 12,000 | 217,900 | 1,092,247,100 | 5,080 |
| Two-wave epidemic[c] | | | | | |
| Vaccine supply 40% | 7,758,800 | 10,600 | 175,100 | 658,380,800 | -- |
| Vaccine supply 20% | 12,765,900 | 19,900 | 371,500 | 702,362,400 | Dominated |
| Vaccine supply 67% | 5,594,000 | 7,800 | 133,700 | 810,775,600 | 3,680 |
| Vaccine supply 80%[b] | 5,940,500 | 6,900 | 119,100 | 891,054,000 | 5,480 |
| No vaccination | 19,290,400 | 70,400 | 1,206,200 | 948,370,400 | Dominated |
| Vaccination pace | | | | | |
| $R_e$ = 1.4 | | | | | |
| Pace 300,000 vaccinations per day | 5,659,400 | 7,200 | 120,300 | 814,977,100 | -- |
| Pace 200,000 vaccinations per day | 8,191,900 | 9,600 | 151,300 | 876,892,500 | Dominated |
| Pace 150,000 vaccinations per day | 10,585,100 | 14,700 | 259,600 | 993,638,400 | Dominated |
| No vaccination | 21,012,100 | 89,300 | 1,558,700 | 1,005,281,800 | Dominated |
| Two-wave epidemic[c] | | | | | |
| Pace 300,000 vaccinations per day | 2,697,100 | 3,200 | 49,300 | 688,458,100 | -- |
| Pace 200,000 vaccinations per day | 4,148,500 | 5,900 | 90,300 | 744,446,000 | Dominated |
| Pace 150,000 vaccinations per day | 5,594,000 | 7,800 | 133,700 | 810,775,600 | Dominated |
| No vaccination | 19,290,400 | 70,400 | 1,206,200 | 948,370,400 | Dominated |

**Supplementary Table 4, continued.**

| Scenario and Vaccination Strategy | Cumulative SARS-CoV-2 infections | Cumulative COVID-19 deaths | Years-of-life lost | Health care costs, USD | ICER, USD per year-of-life saved[a] |
|---|---|---|---|---|---|
| Vaccine supply and vaccination pace | | | | | |
| $R_e$ = 1.4 | | | | | |
| Vaccine supply 40%, pace 300,000 vaccinations per day | 9,866,800 | 13,000 | 211,300 | 688,324,400 | -- |
| Vaccine supply 40%, pace 150,000 vaccinations per day | 11,784,700 | 16,000 | 275,800 | 797,871,600 | Dominated |
| Vaccine supply 67%, pace 300,000 vaccinations per day | 5,659,400 | 7,200 | 120,300 | 814,977,100 | 1,390 |
| Vaccine supply 67%, pace 150,000 vaccinations per day | 10,585,100 | 14,700 | 259,600 | 993,638,400 | Dominated |
| No vaccination | 21,012,100 | 89,300 | 1,558,700 | 1,005,281,800 | Dominated |
| Two-wave epidemic[c] | | | | | |
| Vaccine supply 40%, pace 300,000 vaccinations per day | 6,223,600 | 7,200 | 126,900 | 580,478,200 | -- |
| Vaccine supply 40%, pace 150,000 vaccinations per day | 7,758,800 | 10,600 | 175,100 | 658,380,800 | Dominated |
| Vaccine supply 67%, pace 300,000 vaccinations per day | 2,697,100 | 3,200 | 49,300 | 688,458,100 | 1,390 |
| Vaccine supply 67%, pace 150,000 vaccinations per day | 5,594,000 | 7,800 | 133,700 | 810,775,600 | Dominated |
| No vaccination | 19,290,400 | 70,400 | 1,206,200 | 948,370,400 | Dominated |

ICU: intensive care unit. USD: United States dollars. ICER: incremental cost-effectiveness ratio. $R_e$: effective reproduction number. Dominated: the strategy results in a higher ICER than that of a more clinically effective strategy, or the strategy results in less clinical benefit (more years-of-life lost) and higher health care costs than an alternative strategy.

In this analysis, daily costs of hospital care ($137) and ICU care ($798) were applied from a report by Edoka et al.[3] These values did not include facility costs.

[a]Within each $R_e$ scenario, vaccination strategies are ordered from lowest to highest cost per convention of cost-effectiveness analysis. ICERs are calculated compared to the next least expensive, non-dominated strategy. Displayed life-years and costs are rounded to the nearest hundred, while ICERs are calculated based on non-rounded life-years and costs and then rounded to the nearest ten.

[b]When modeling a vaccination program that seeks to vaccinate 80% of the population, uptake among those eligible was increased to 80% to avoid a scenario in which supply exceeds uptake. If uptake is not increased beyond 67%, then the strategy of vaccinating 67% of the population provides the most clinical benefit and results in an ICER of $12,110/YLS compared with vaccinating 40% of the population when $R_e$ is 1.4 and $3,680/YLS in an epidemic scenario with periodic surges.

[c]In the analysis of an epidemic with periodic surges, the basic reproduction number ($R_o$) alternates between low and high values over time, and the $R_e$ changes day-to-day as the epidemic and vaccination program progress and there are fewer susceptible individuals. For most of the simulation horizon, $R_o$ is 1.6 (equivalent to an initial $R_e$ of 1.1). However, during days 90-150 and 240-300 of the simulation, $R_o$ is increased to 2.6. This results in two epidemic waves with peak $R_e$ of approximately 1.4-1.5.

**Supplementary Table 5. Additional model input parameters.**

| Parameter | Base case value (Range) | Sources |
|---|---|---|
| Initial state | | |
| Initial disease distribution | | |
| Susceptible | 69.9 (49.9-89.9) | Assumption |
| Pre-infectious incubation | 0.015 | Assumption |
| Asymptomatic | 0.040 | Assumption |
| Mild or moderate disease | 0.045 | Assumption |
| Severe disease | 0.00005 | Assumption |
| Critical disease | 0.00005 | Assumption |
| Recuperation after critical disease | 0.0 | Assumption |
| Recovered (prior immunity) | 30 (10-50) | [4–7] |
| Transmission dynamics | | |
| Rate of onward transmission by health state[a], transmissions per person-day | | [8–12] |
| Asymptomatic | 0.1858 | |
| Mild or moderate disease | 0.1512 | |
| Severe disease | 0.0106 | |
| Critical disease | 0.0084 | |
| Recuperation after critical disease | 0.0106 | |

**Supplementary Table 5, continued. Additional model input parameters.**

| Parameter | Base case value (Range) | | | | Sources |
|---|---|---|---|---|---|
| Natural history of COVID-19 disease | | | | | |
| Disease path probability[b], stratified by age, % | Asymp. | Mild / Mod | Severe | Critical | [8,13–16] |
| Ages <20 years | 29.93 | 69.78 | 0.25 | 0.03 | |
| Ages 20-59 years | 17.90 | 80.38 | 0.80 | 0.93 | |
| Ages ≥60 years | 17.10 | 76.37 | 1.36 | 5.16 | |
| Daily disease progression probability, stratified by path, % | Asymp. | Mild / Mod. | Severe | Critical | [8–11,17] |
| From pre-infectious incubation to asymptomatic | 31.93 | 31.93 | 31.93 | 31.93 | |
| From asymptomatic to mild or moderate disease | -- | 39.35 | 39.35 | 39.35 | |
| From mild or moderate disease to severe disease | -- | -- | 14.26 | 28.35 | |
| From severe disease to critical disease | -- | -- | -- | 20.17 | |
| From critical disease to recuperation | -- | -- | -- | 9.61[c] | |
| Daily recovery probability, stratified by path, % | Asymp. | Mild / Mod. | Severe | Critical | [8–11,17] |
| From asymptomatic | 9.99 | -- | -- | -- | |
| From mild or moderate disease | -- | 9.52 | -- | -- | |
| From severe disease | -- | -- | 9.08 | 5.48[d] | |
| From recuperation after critical disease | -- | -- | -- | 16.09 | |
| Daily mortality probability while critical, stratified by age, % | Ages <20 years | Ages 20-59 years | Ages ≥60 years | | |
| Without ICU care | 11.75 | 16.62 | 20.33 | | Assumption |
| With ICU care | 0.0006 | 0.38 | 5.00 | | [8–11,17] |

Asymp: Asymptomatic. Mod: Moderate. ICU: Intensive care unit.

[a]This corresponds to $R_e$ of 1.4.

[b]Disease path probabilities are for unvaccinated individuals susceptible to SARS-CoV-2 infection.

[c]The transition from the critical state to recuperation requires admission to an intensive care unit (ICU).

[d]Early recovery from the severe state on the critical path requires admission to a hospital.

### Supplementary Methods

#### Overview

The Clinical and Economic Analysis of COVID-19 Interventions (CEACOV) model incorporates aspects of SARS-CoV-2 transmission, COVID-19 disease natural history, and vaccination to project epidemic growth, clinical outcomes, health care resource utilization, and costs.

Projections for each scenario examined are generated by simulating a cohort of 1 million individuals over a 360-day time horizon. Simulation outputs are subsequently scaled to reflect the mid-year 2019 population of South Africa (approximately 59 million people) to produce country-level estimates of clinical and economic outcomes.[18] We measured the clinical benefit of a vaccination program in terms of the undiscounted years-of-life saved (YLS) from averted COVID-19 deaths. We considered health care costs associated with vaccination, hospital care, and intensive care unit (ICU) care from an all-payer (public and private) health sector perspective.

We calculated the incremental cost-effectiveness ratio (ICER) of a vaccination program. In comparisons of different levels of vaccine supply and vaccination pace, the ICER of a given strategy was calculated by dividing the change in costs by the change in clinical benefits (years-of-life) compared with the next least-expensive, non-dominated strategy. A strategy is ruled out though strong dominance if it is more expensive and less effective (resulting in more years-of-life lost) than an alternative strategy, or through extended dominance if it is less economically efficient (resulting in a higher ICER) than a more effective strategy.[19] In the base case and sensitivity analyses, in which we varied factors that are less in the direct control of policymakers, such as vaccine characteristics and epidemic growth, we compared the ICER of a reference vaccination program (supply sufficient to vaccinate 67% of South Africa's population, pace 150,000 vaccinations per day) with a scenario without vaccines.

**Health states**

CEACOV is a dynamic microsimulation model of COVID-19 based on a stochastic susceptible-exposed-infectious-recovered (SEIR) framework. Individuals face daily state-transition probabilities that govern movement between the following health states (Supplementary Table 5, Supplementary Fig. 1):

- *Susceptible*: individuals who are not currently infected with SARS-CoV-2 but may become infected during the simulation
- *Pre-infectious incubation*: individuals who have been exposed to SARS-CoV-2 but are not yet infectious to others
- *Asymptomatic*: individuals infected with SARS-CoV-2 who do not display symptoms but can transmit the virus to others
- *Mild/Moderate*: individuals infected with SARS-CoV-2 who display mild/moderate symptoms but do not require hospitalization
- *Severe*: individuals with severe symptoms of COVID-19 ideally managed in a hospital but not requiring admission to an ICU
- *Critical*: individuals with life-threatening symptoms and signs of COVID-19 who require ICU care to survive
- *Recuperation*: individuals recuperating from critical illness while remaining in the hospital and can still transmit the virus to others
- *Recovered*: individuals fully recovered from COVID-19, who can no longer transmit the virus to others and are immune from reinfection for the duration of the simulation period
- *Dead*: individuals who die as a result of critical COVID-19 disease

The state transition from susceptible to infected health states depends on the prevalence of active COVID-19 disease at a given point in time. Once infected, individuals have an age-dependent probability of progressing along one of four "paths" culminating in either asymptomatic infection, mild/moderate disease, severe disease, or critical disease (Supplementary Table 5, Supplementary Fig. 1). Before reaching a more advanced disease state, individuals must first transition through intermediate states (e.g., individuals destined for severe disease must first pass through the asymptomatic and mild/moderate health states).

Hospital and ICU care reduce the probability of death for those on the critical disease path in two ways: by providing a chance at early recovery to those in the severe state who have access to a hospital bed (allowing them to bypass the critical state) and by increasing the chances of survival for those in the critical state who have access to ICU care. The CEACOV model incorporates constraints on health care resources, and in the event hospital or ICU capacity is reached, individuals default to the next highest level of treatment.

**Prevalence of prior immunity**

We assumed that those infected by SARS-CoV-2 prior to the emergence of the B.1.351 variant were susceptible to reinfection and those who infected after the emergence of B.1.351 were immune from reinfection for the model simulation period. We used the following approach to estimate the cumulative number of people in South Africa who were infected with the B.1.351 variant by 30 January 2021:

1) We assumed that 100% of infections occurring after 14 November 2020 were attributable to the B.1.351 variant. An analysis of genomic surveillance data suggested that by the end of November 2020, this variant accounted for more than 75% of new infections in four provinces.[4]

2) We compared the number of cases reported to South Africa's National Institute for Communicable Diseases between 14 November 2020 and 30 January 2021 (all assumed to be attributable to the B.1.351 variant) to the number of cases reported before 14 November 2020 (all assumed to be attributable to ancestral variants). In this manner, we estimated the proportion of cumulative infections attributable to B.1.351 in Eastern Cape, Free State, KwaZulu-Natal, and Northern Cape provinces.[6,7]
3) For each province, we multiplied the prevalence of SARS-CoV-2 seropositivity among blood donors in January 2021 by the estimated proportion of cumulative infections attributable to B.1.351 to derive the prevalence of prior infection by B.1.351.[5]
4) Using weighted averages based on the population of each province, we estimated the proportion of people across South Africa who have been previously infected by B.1.351.[18]

From this approach and rounding, we estimated that 30% of individuals have previously been infected with the B.1.351 variant and are thus immune from reinfection for the simulation period. To account for uncertainty in our estimates, we varied the prevalence of prior immunity between 10% and 50% in sensitivity analyses.

**Transmission**

Simulated individuals in asymptomatic, mild/moderate, severe, critical, or recuperation states of COVID-19 can transmit infection to susceptible individuals at state-dependent daily rates. The number of daily infections is determined by the proportion of susceptible people in the population and the distribution of disease states among those with COVID-19. Our model assumes that the force of infection is the same for all age groups (i.e., homogeneous mixing). This assumption is supported by two large seroprevalence studies in South Africa, which both demonstrated similar seroprevalence within age bands, both with community-based testing and with testing of blood donors.[5,20]

As described previously, we calibrated transmission rates to correspond to a particular initial effective reproduction number ($R_e$), which was 1.4 in the base case of this analysis.[21,22] In sensitivity analysis, we varied the initial $R_e$ by changing the transmission rates. We also evaluated a scenario in which there were two surges in infections. We did this by alternating the basic reproduction number ($R_o$) between low and high values over time. This results in the $R_e$ changing day-to-day as the epidemic and vaccination program progress and there are fewer susceptible individuals. For most of the simulation horizon, $R_o$ is 1.6 (equivalent to an initial $R_e$ of 1.1). However, during days 90-150 and 240-300 of the simulation, $R_o$ is increased to 2.6. This results in two epidemic waves with peak $R_e$ of approximately 1.4-1.5 (Figure S10).

**Disease natural history**

Once infected with SARS-CoV-2, an individual moves to the pre-infectious latency state and faces age-dependent probabilities of developing asymptomatic, mild/moderate, severe, or critical disease (Supplementary Table 5, Supplementary Figure 1). Those with critical disease face age-dependent daily probabilities of death. If they survive, they transition to a recuperation state (remaining infectious) before going to the recovery (immune) state. Those in other disease states can transition directly to the recovery state. “Recovered” individuals are assumed immune from repeat infection for the simulation duration and pose no risk of transmission. To capture infection transmission dynamics, all simulated individuals advance through the model simultaneously.

We calculated age-stratified disease path probabilities based on data indicating the proportion of people in each age group who developed asymptomatic infection, disease requiring hospitalization, and disease requiring ICU care.[8,13–16] We derived the average duration that infected individuals spend in each health state based on the following sources: the pre-infectious times reported by the WHO-China Joint Mission on COVID-19[8] and He et al.;[9] time to development of pneumonia (Wang et al.[17]); time to ICU admission and time spent in the ICU (Zhou et al.[10]); and median time to death (Zhou et al.[10]). We derived the time to recovery from the durations of viral shedding based on polymerase chain reaction (PCR) detectability.[8,10,11] After determining the average duration of each health state (measured in days), we calculated state transition rates as follows:

$$\text{Transition rate} = r = (\text{average duration of health state})^{-1} \tag{1}$$

Based on the transition rates estimated using equation (1), we then calculated daily state-transition probabilities as:

$$\text{Daily transition probability} = 1 - exp(-r\Delta t) \tag{2}$$

Where $\Delta t$ represents the chosen time step duration – in this case, one day.

**Life expectancy and years-of-life lost**

We quantified the clinical benefit of a vaccination program in terms of years-of-life saved (YLS) from averted COVID-19 deaths. To do this, we calculated the number of years a person would have lived subsequent to their current age had they not died of COVID-19. Our approach has been detailed elsewhere.[22]

Since our model uses large age groups (<20 years, 20-59 years, ≥60 years), we calculated the average years-of-life lost per COVID-19 death in each of our model's age groups ($\overline{YLL}$) by calculating a weighted average of remaining life expectancy in five-year intervals over the age distribution of deaths within each large age group:

$$\overline{YLL} = \frac{\sum_i w_i LE_i}{\sum_i w_i} \tag{3}$$

where $w_i$ represents the relative frequency at which someone age $i$ is expected to die from COVID-19, and $LE_i$ represents the remaining life expectancy among those age $i$. The age-stratified distribution of COVID-19 deaths was determined from data reported by South Africa's Department of Health.[23] Age-stratified life expectancy was calculated by generating a standard abridged life table for South Africa's population using the method described by Preston et al.[24] Mortality trends were derived from estimates of all-cause mortality for the year 2016 published by the World Health Organization and age/sex-stratified population estimates published by the United Nations World Population Prospects.[25,26]

**Vaccine effectiveness and outcomes**

To reconcile the various vaccine efficacy endpoints used in vaccine clinical trials, we incorporated three measures of vaccine effectiveness into our model: effectiveness in preventing SARS-CoV-2 infection ($VE_I$), effectiveness in preventing mild/moderate COVID-19 disease ($VE_M$), and effectiveness in preventing severe or critical COVID-19 disease that would prompt hospitalization ($VE_H$). Severe or critical COVID-19 disease was defined as illness that would ordinarily prompt non-ICU hospital care or ICU care, respectively. Of note, these definitions applied in our model are somewhat different from the definitions applied in various clinical trials.[27–30]

We accounted for the direct benefits of vaccination by adjusting disease path probabilities among those vaccinated. Disease path probabilities are age-dependent and defined as follows:

- $P_0$: probability immune to SARS-CoV-2 infection

- $P_1$: probability of developing asymptomatic SARS-CoV-2 infection if exposed
- $P_2$: probability of developing mild/moderate COVID-19 disease if exposed
- $P_3$: probability of developing severe COVID-19 disease if exposed
- $P_4$: probability of developing critical COVID-19 disease if exposed

Disease path probabilities for unvaccinated individuals susceptible to SARS-CoV-2 infection are shown in Supplementary Table 5. Among vaccinated individuals exposed to SARS-CoV-2, the probabilities of infection, mild/moderate disease, severe disease, or critical disease are functions of vaccine effectiveness and disease path probabilities among unvaccinated individuals exposed to SARS-CoV-2:

$$P(\text{immune}) = P_0^V = 1 - \big((1 - VE_I)(1 - P_0)\big) \quad (4)$$

$$P(\text{asymptomatic infection}) = P_1^V = 1 - (P_0^V + P_2^V + P_3^V + P_4^V) \quad (5)$$

$$P(\text{mild/moderate disease}) = P_2^V = (1 - VE_M)P_2 \quad (6)$$

$$P(\text{severe disease}) = P_3^V = (1 - VE_H)P_3 \quad (7)$$

$$P(\text{critical disease}) = P_4^V = (1 - VE_H)P_4 \quad (8)$$

Supplementary Figure 2 depicts a graphical summary of the above equations. In the base case, we assumed that those who are vaccinated but nonetheless become infected can transmit infection to susceptible people with the same probability as unvaccinated/infected people can transmit infection. In a sensitivity analysis, we reduced the probability of transmission from vaccinated/infected people by 50%.

Our modeled vaccine effectiveness values were based on efficacies reported in the Johnson & Johnson/Janssen Ad26.COV2.S vaccine trial.[30] Of note, because of slightly differing outcome definitions, our modeled vaccine effectiveness in preventing mild/moderate COVID-19 was adjusted in an age-dependent manner to reflect the trial's reported efficacy in preventing moderate to severe/critical COVID-19 in South Africa, and our modeled vaccine effectiveness in preventing severe/critical COVID-19 that would prompt hospitalization reflected the trial's reported efficacy in preventing COVID-19 requiring medical intervention.[30]

**Costs of vaccination, hospital ward care, and ICU care**

Although we did not perform a full economic analysis of vaccination program costs, we accounted for the estimated cost of each vaccine dose and of vaccine delivery in South Africa. We applied a reported cost of US$10 per dose for the single-dose Johnson & Johnson/Janssen Ad26.COV2.S vaccine in South Africa.[31,32] We applied a service and delivery cost of US$4.81 per dose, based on a published report of the costs of delivering human papillomavirus (HPV) vaccination in South Africa.[33] In that report, the authors found that service and delivery costs for a 3-dose regimen of HPV vaccine were 180.28 South African rand (ZAR) per person (in 2014 ZAR). Converting from 2014 ZAR to 2020 ZAR, and from 2020 ZAR to 2020 US dollars, yields a cost of US$14.42 per person.[34,35] Dividing by three results in a cost of US$4.81 per dose.

In the base case, we applied a daily cost of $154 for hospital (non-ICU) ward care based on cost per inpatient day in a general ward charged by Netcare, a private hospital network.[36] Those in the "severe" and "recuperation" states accrued this daily cost, assuming a hospital ward bed was available. In the base case, we applied a daily cost of $1,751 for ICU care based on a cost analysis of a public hospital in KwaZulu-Natal, South Africa.[37] Those in the "critical" state accrued this daily cost and not the hospital ward care cost, assuming an ICU bed was available. We performed sensitivity analysis in which we varied the hospital and ICU care costs to 50% and 200% of the base case values.

Additionally, we repeated the main analysis using costs reported by Edoka et al. in the public sector.[3] These were $137/day for hospital (non-ICU) general ward care with supplemental oxygen and $798/day for ICU care with invasive mechanical ventilation, both excluding facility fees (when including facility fees, Edoka et al. reported costs of $202/day for hospital ward care and $1,314/day for ICU care). The results of this analysis are in Supplementary Table 4.

**Model validation**

We previously validated CEACOV model results by comparing model-projected COVID-19 deaths with those reported in South Africa.[22] Here, we performed additional validation by simulating a two-wave epidemic, starting with 0.1% prevalence of active SARS-CoV-2 infection and 0% prevalence of prior immunity (analogous to South Africa in early 2020) (Figure S3). We compared the model results to COVID-19 case and death data reported to South Africa's National Institute for Communicable Diseases, assuming that reported cases represent 10% of all infections.[1,2] Our model projected, between 1 May 2020 and 10 April 2021, 15.6 million SARS-CoV-2 infections, 1,555,900 observed cases, and 58,200 COVID-19 deaths. During this same period, 1,551,768 cases of COVID-19 were reported to the NICD. Weekly data regarding the number of reported deaths were unavailable before 18 May 2020. However, between 5 March 2020 and 10 April 2021, 52,278 COVID-19 deaths were reported to the NICD. These data suggest that our model is able to capture the general characteristics of South Africa's COVID-19 epidemic.
